# Prevalence of Tobacco Smoking and Associated Factors Among High School Students in Harar Town, Eastern Ethiopia: A School-Based Cross-Sectional Study

**DOI:** 10.64898/2026.09.30.26364450

**Authors:** Ephrem Zeleke Detamo, Mekonnen Sisay, Tariku Dingeta Amante

## Abstract

**Background:** Tobacco use during adolescence is an important public-health concern because early initiation increases the likelihood of continued tobacco use and dependence. This study assessed the prevalence of tobacco smoking and factors associated with ever smoking among high school students in Harar town, eastern Ethiopia.

**Methods:** A school-based cross-sectional study was conducted among 467 students in grades 9–12 from selected public and private high schools in Harar town. Data were collected using a self-administered questionnaire adapted from the Global Youth Tobacco Survey. Descriptive statistics and binary logistic regression were used. Variables were entered into a multivariable logistic regression model based on the prespecified analytical approach, and adjusted odds ratios (AORs) with 95% confidence intervals (CIs) were estimated. Statistical significance was declared at p<0.05.

**Results:** Among 467 students, 64 (13.7%; 95% CI 10.9–17.1%) reported ever smoking tobacco, while 25 (5.4%; 95% CI 3.7–7.8%) were current smokers. In the SPSS multivariable logistic regression model, grade 11 (AOR 10.61, 95% CI 3.52–31.97) and grade 12 (AOR 12.18, 95% CI 3.95–37.53), alcohol drinking (AOR 2.60, 95% CI 1.25–5.40), khat chewing (AOR 9.57, 95% CI 4.04–22.65), and access to tobacco (AOR 3.18, 95% CI 1.58–6.43) were independently associated with ever smoking. Male sex, age ≥18 years, and grade 10 were not statistically significant in the adjusted model.

**Conclusion:** Ever smoking was reported by approximately one in seven students and current smoking by about one in twenty. Alcohol drinking, khat chewing, access to tobacco, and higher grade were independently associated with ever smoking. School- and community-based tobacco prevention should address substance use and the availability of tobacco products around schools.

## Introduction

Tobacco use remains a major preventable cause of morbidity and mortality worldwide. Adolescence is a critical period for tobacco prevention because experimentation during the school years can progress to regular use and dependence. The burden is particularly important in low- and middle-income countries, where tobacco control measures may be incompletely implemented and social and environmental influences can facilitate access to tobacco products.

Ethiopian studies have documented substantial variation in adolescent tobacco use across settings. Previous work in eastern Ethiopia reported a cigarette-smoking prevalence of 12.2% among high school students. Other Ethiopian and international studies have identified age, sex, substance use, peer and family influences, and access to tobacco as potential determinants. However, evidence from Harar town remains limited, particularly regarding factors associated with tobacco smoking beyond cigarettes.

This study therefore assessed the prevalence of tobacco smoking and factors associated with ever smoking among high school students in Harar town, eastern Ethiopia.

## Methods

### Study design and setting

A school-based cross-sectional study was conducted in Harar town, eastern Ethiopia, from January 4 to February 26, 2021. Harar is the capital of Harari Regional State and is located approximately 526 km east of Addis Ababa. At the time of the study, the town had 14 high schools (9 public and 5 private). Four schools—two public and two private—were selected for the study.

### Population and sampling

The source population comprised regular students in grades 9–12 attending public and private high schools in Harar town. Students who were seriously ill and unable to respond during data collection were excluded. Two public and two private schools were selected using simple random sampling, followed by proportional allocation by grade and random selection from the student sampling frames. The final analytic dataset contained 467 complete records.

### Sample size

The original manuscript reported a target sample of 495 based on an anticipated prevalence of 12.2%, a 95% confidence level, 3% precision, and 10% non-response. Recalculation shows that the stated assumptions yield approximately 458 participants before allowance for non-response and approximately 509 when the non-response allowance is applied by division by 0.90. Therefore, the previously reported value of 495 is not reproducible from the stated assumptions. The present analysis uses all 467 complete records in the final dataset and does not claim that 495 is mathematically derived from those assumptions.

### Data collection and quality control

Data were collected using a structured, self-administered questionnaire adapted from the Global Youth Tobacco Survey and modified to the Ethiopian context. The questionnaire was translated into Amharic and Afaan Oromo and pretested before the survey. Four teachers served as data collectors and were assigned outside their usual schools to reduce social-desirability bias. One supervisor from the Harari Regional Education Bureau supervised fieldwork. Questionnaires were checked for completeness and consistency, and data were double-entered in EpiData before export to SPSS. The final CSV used for this report contains 467 records and 57 variables.

### Ethical-considerations

Ethical clearance was obtained from the Haramaya University Health Research Ethics Review Committee (IHRERC), approval/reference number IHRERC/001/2021. Permission was obtained from the Harari Regional Education Bureau and participating schools. Data collection was conducted from January 1 to February 26, 2021. Written informed consent was obtained from participants. For students younger than 18 years, written informed consent was obtained from their parents or guardians before data collection. Participation was voluntary, confidentiality was maintained, and names or school identification numbers were not recorded on the questionnaires.

### Variables and operational definitions

The primary outcome was ever tobacco smoking, defined as having ever tried tobacco smoking, including one or two puffs. Current smoking was defined as smoking any tobacco product on one or more days during the 30 days preceding the survey. The principal explanatory variables included age, sex, grade, alcohol drinking, khat chewing, and access to tobacco products. Tobacco smoking included cigarettes and other smoked tobacco products such as shisha, cigars and Gaya, consistent with the study protocol.

### Statistical analysis

The final SPSS dataset and the accompanying SPSS output were reviewed. Descriptive statistics were summarized using frequencies and percentages. Binary logistic regression was used with ever smoking as the dependent variable. Crude odds ratios (CORs) and 95% CIs were estimated for candidate variables. Variables selected according to the prespecified analytical approach were entered simultaneously into a multivariable logistic regression model. The final SPSS model included nine predictor blocks: Rec_DOUCANYHLZWAR, Rec_BUYNEAR2SCHOOL, sex, age, khat chewing, alcohol drinking, Rec_LRNABTTOBACP12, grade, and Rec_DAN_TOBACC_SMG. Adjusted odds ratios (AORs), 95% CIs and p-values were taken directly from the SPSS output. Statistical significance was defined as p<0.05.

The multivariable results were reconciled against the newly located SPSS output document. The SPSS output provides the complete final model, including the omnibus model test, Hosmer–Lemeshow test, all coefficients, confidence intervals, and the variables entered at Step 1. Where the CSV export did not preserve human-readable value labels, the original SPSS variable names are retained in the reconciliation table rather than assigning unsupported labels.

## Results

### Socio-demographic characteristics

| Variable | Category | n | % |
| --- | --- | --- | --- |
| Sex | Male | 235 | 50.3 |
|  | Female | 232 | 49.7 |
| Age | <18 years | 229 | 49.0 |
|  | ≥18 years | 238 | 51.0 |
| Religion | Orthodox | 220 | 47.1 |
|  | Muslim | 152 | 32.5 |
|  | Protestant | 83 | 17.8 |
|  | Catholic | 6 | 1.3 |
|  | Other | 6 | 1.3 |
| Grade | 9 | 173 | 37.0 |
|  | 10 | 108 | 23.1 |
|  | 11 | 85 | 18.2 |
|  | 12 | 101 | 21.6 |
| Mother education | Illiterate | 45 | 9.6 |
|  | Read & write | 64 | 13.7 |
|  | Grades 1–8 | 89 | 19.1 |
|  | Grades 9–12 | 114 | 24.4 |
|  | Diploma or above | 155 | 33.2 |
| Father education | Illiterate | 34 | 7.3 |
|  | Read & write | 41 | 8.8 |
|  | Grades 1–8 | 46 | 9.9 |
|  | Grades 9–12 | 101 | 21.6 |
|  | Diploma or above | 245 | 52.5 |
| Pocket money | Never | 55 | 11.8 |
|  | Few times | 205 | 43.9 |
|  | Once a month | 66 | 14.1 |
|  | More than once a month | 141 | 30.2 |

Note: The frequencies above are recalculated directly from the final CSV. They replace discrepant frequencies in the earlier manuscript, particularly the religion and grade distributions.

### Prevalence of tobacco smoking

| Outcome | Category | n | % |
| --- | --- | --- | --- |
| Ever smoking | No | 403 | 86.3 |
|  | Yes | 64 | 13.7 |
| Current smoking | No | 442 | 94.6 |
|  | Yes | 25 | 5.4 |
| Other tobacco smoking | No | 435 | 93.1 |
|  | Yes | 32 | 6.9 |

### Passive smoking and tobacco-related attitudes

| Variable | Category | n | % |
| --- | --- | --- | --- |
| Household smoking in previous 7 days | 0 days | 355 | 76.0 |
|  | 1–2 days | 57 | 12.2 |
|  | 3–4 days | 29 | 6.2 |
|  | 5–6 days | 11 | 2.4 |
|  | 7 days | 15 | 3.2 |
| Smoking inside enclosed public places | 0 days | 280 | 60.0 |
|  | 1–2 days | 89 | 19.1 |
|  | 3–4 days | 55 | 11.8 |
|  | 5–6 days | 24 | 5.1 |
|  | 7 days | 19 | 4.1 |
| Smoking at outdoor public places | 0 days | 268 | 57.4 |
|  | 1–2 days | 84 | 18.0 |
|  | 3–4 days | 64 | 13.7 |
|  | 5–6 days | 31 | 6.6 |
|  | 7 days | 20 | 4.3 |
| Smoking on school property | No | 331 | 70.9 |
|  | Yes | 136 | 29.1 |
| Support for banning smoking in enclosed public places | Yes | 357 | 76.4 |
|  | No | 110 | 23.6 |
| Support for banning smoking at outdoor public places | Yes | 352 | 75.4 |
|  | No | 115 | 24.6 |

### Access to tobacco and tobacco-control information

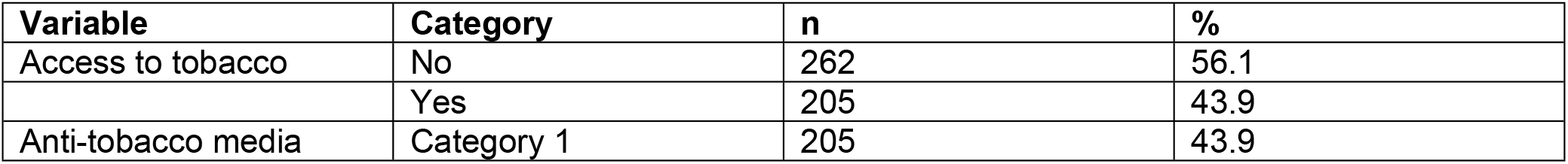
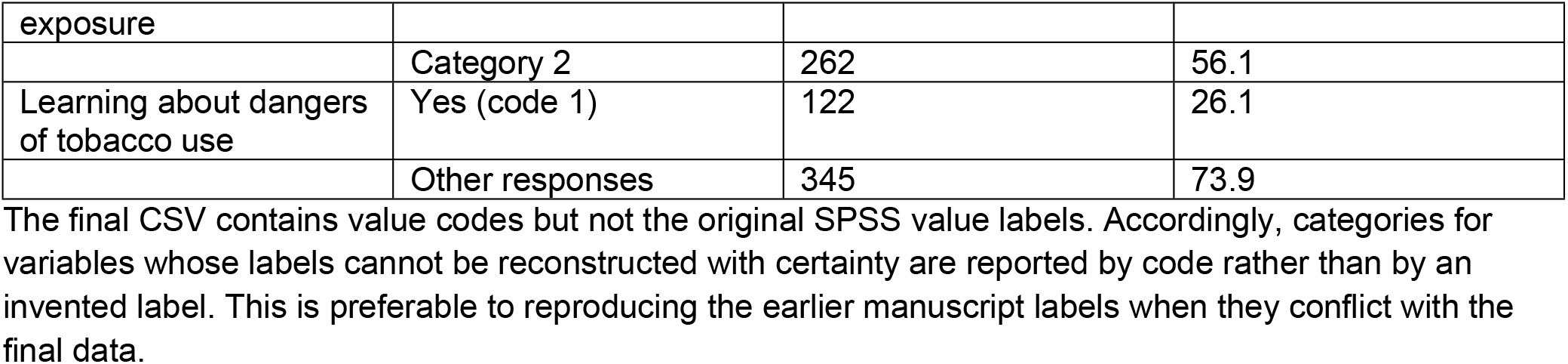

### Factors associated with ever smoking

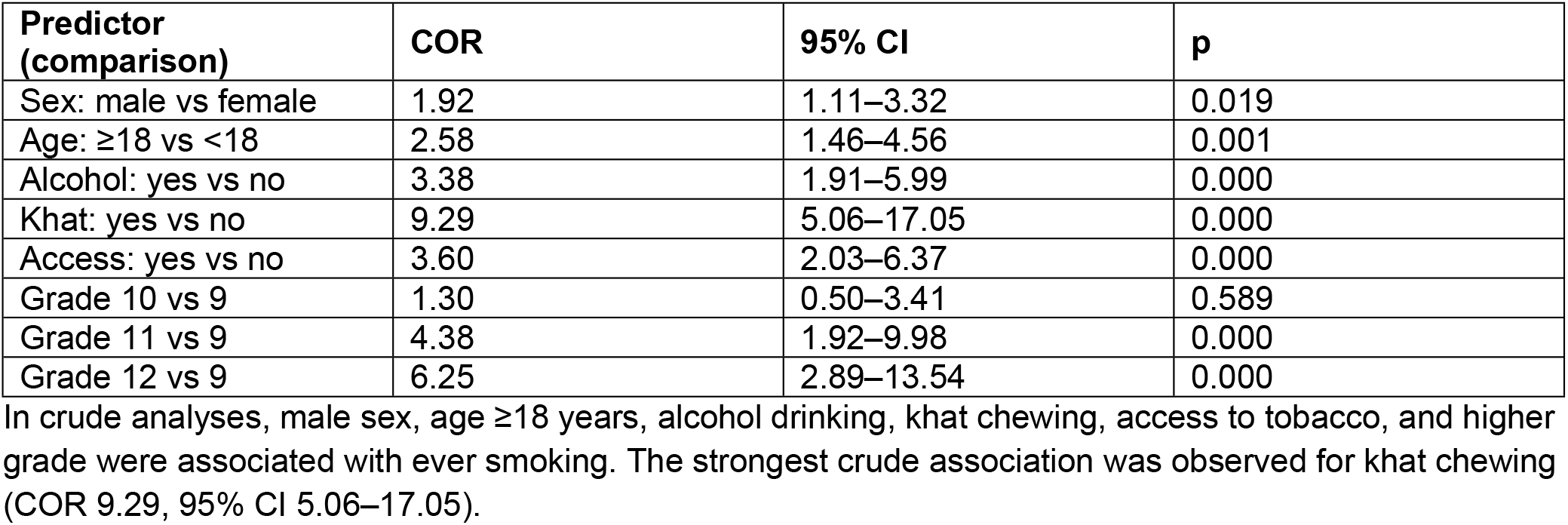

### Multivariable logistic regression

| Predictor (comparison) | AOR | 95% CI | p |
| --- | --- | --- | --- |
| Rec_DOUCANYHLZWAR (1) | 1.099 | 0.557–2.168 | 0.786 |
| Access to tobacco: yes vs no | 3.182 | 1.576–6.427 | 0.001 |
| Male vs female | 1.059 | 0.514–2.183 | 0.877 |
| Age ≥18 vs <18 years | 0.838 | 0.364–1.928 | 0.678 |
| Khat chewing: yes vs no | 9.566 | 4.041–22.645 | <0.001 |
| Alcohol drinking: yes vs no | 2.599 | 1.250–5.403 | 0.011 |
| Rec_LRNABTTOBACP12 (1) | 0.676 | 0.321–1.424 | 0.303 |
| Grade 10 vs grade 9 | 2.107 | 0.636–6.977 | 0.223 |
| Grade 11 vs grade 9 | 10.610 | 3.521–31.968 | <0.001 |
| Grade 12 vs grade 9 | 12.180 | 3.952–37.533 | <0.001 |
| Rec_DAN_TOBACC_SMG (1) | 4.255 | 1.465–12.358 | 0.008 |
| Rec_DAN_TOBACC_SMG (2) | 8.977 | 2.156–37.371 | 0.003 |
| Rec_DAN_TOBACC_SMG (3) | 1.157 | 0.525–2.548 | 0.718 |

Model fit: n=467; likelihood-ratio test $p < 0.001$ ; McFadden pseudo- $R^2 = 0.303$ .

After adjustment, grade 11 and grade 12, alcohol drinking, khat chewing, and access to tobacco remained independently associated with ever smoking. Compared with grade 9, the adjusted odds were 10.61-fold higher in grade 11 and 12.18-fold higher in grade 12. Students reporting alcohol drinking had 2.60 times the adjusted odds of ever smoking, those reporting khat chewing had 9.57 times the adjusted odds, and those reporting access to tobacco had 3.18 times the adjusted odds. Male sex, age ≥18 years, and grade 10 were not statistically significant after adjustment. The overall model was statistically significant (likelihood-ratio χ^2^=130.459, df=13, p<0.001), and the Hosmer–Lemeshow test was not statistically significant (χ^2^=17.318, df=8, p=0.270).

## Discussion

This study found that 13.7% of high school students had ever smoked tobacco and 5.4% were current smokers. The findings indicate that tobacco experimentation remains common among adolescents in Harar town, even though current smoking was less frequent than ever smoking. The distinction between ever smoking and current smoking is important and should be maintained throughout interpretation.

Higher grade was strongly associated with ever smoking after adjustment. The progressively higher odds in grades 11 and 12 may reflect increasing age, greater independence, longer exposure to peer and community influences, and greater opportunity to access tobacco products. Because this was a cross-sectional study, however, the association should not be interpreted as causal.

Alcohol drinking and khat chewing were independently associated with ever smoking. The co-occurrence of substance-use behaviors among adolescents may reflect shared social environments, peer influences, or behavioral clustering. Prevention programs should therefore avoid addressing tobacco in isolation and should incorporate broader substance-use prevention and risk communication.

Access to tobacco was also independently associated with ever smoking. Easy availability of tobacco products may lower barriers to experimentation. Strengthening enforcement of restrictions on tobacco sales around schools and improving school-community tobacco-control measures may therefore be useful components of prevention.

The dataset also documents substantial exposure to second-hand smoke. A sizeable proportion of students reported exposure in households, enclosed public places, and outdoor public places, and 29.1% reported smoking inside the school building or on school property. These findings support a comprehensive tobacco-control approach that combines prevention of initiation with protection from second-hand smoke.

## Strengths and limitations

A strength of this study is the use of a school-based sample from both public and private schools and a structured questionnaire adapted from the Global Youth Tobacco Survey. The present analysis also used the complete final dataset and recalculated the reported frequencies and regression estimates rather than relying on previously published tables.

The study has several limitations. Its cross-sectional design prevents temporal or causal inference. Tobacco use was self-reported and may be affected by recall or social-desirability bias. The relatively small number of ever smokers (64) limits precision for multivariable estimates, particularly for categorical predictors with several levels. In addition, several predictor labels in the CSV export were represented only by recoded variable names; these have therefore been retained exactly as they appear in the original SPSS output where a human-readable label could not be independently verified.

## Conclusion

Ever tobacco smoking was reported by 13.7% of high school students in Harar town, while 5.4% were current smokers. Higher grade, alcohol drinking, khat chewing, and access to tobacco were independently associated with ever smoking. Tobacco prevention efforts in Harar should combine school-based education with stronger control of tobacco availability and integrated prevention of alcohol and khat use. Protection from second-hand smoke should also remain a priority.

### What this study adds

This study provides updated, dataset-verified estimates of tobacco smoking among high school students in Harar town and demonstrates independent associations between ever smoking and higher grade, alcohol drinking, khat chewing, and access to tobacco products.

## Data Availability

The data underlying the findings of this study contain information collected from school-going adolescents and are subject to ethical considerations regarding participant confidentiality. The data cannot be made publicly available in a manner that could compromise participant privacy. A de-identified dataset may be made available to qualified researchers upon reasonable request, subject to approval by the relevant institutional ethics committee and applicable ethical and data-protection requirements.

## Declarations

### Funding

[CONFIRM BEFORE SUBMISSION: state whether the authors received any funding for this study.]

Ethics approval and consent: Ethical clearance was obtained from Haramaya University Health Research Ethics Review Committee (IHRERC), approval/reference number IHRERC/001/2021 (to be verified against the original clearance letter before submission). Permission was obtained from the Harari Regional Education Bureau and participating schools. Written informed consent was obtained from participants; for students younger than 18 years, written informed consent was obtained from their parents/guardians before data collection. Participation was voluntary, confidentiality was maintained, and names or school identification numbers were not recorded on questionnaires.

### Authors’ contributions

EZ coordinated the study, data collection, analysis and manuscript preparation. MS and TD contributed to study implementation, supervision and manuscript review, consistent with the original manuscript.

### Data availability

Because the study involved human participants who were high-school students, the individual-level dataset is not publicly posted. De-identified data may be made available upon reasonable request, subject to approval under the applicable ethical and institutional requirements. Requests should be directed to the Haramaya University Health Research Ethics Review Committee (IHRERC), Office Phone: 0254662011, P.O. Box 235, Harar, Ethiopia.

## Statistical reconciliation note

The newly located multivariable SPSS output resolves the previous uncertainty. The reported adjusted odds ratios in this final manuscript are taken directly from the final SPSS multivariable model. The model was statistically significant (Omnibus test χ^2^=130.459, df=13, p<0.001), and the Hosmer–Lemeshow goodness-of-fit test was not statistically significant (χ^2^=17.318, df=8, p=0.270), supporting adequate model fit.

## Final multivariable logistic regression model fit (SPSS)

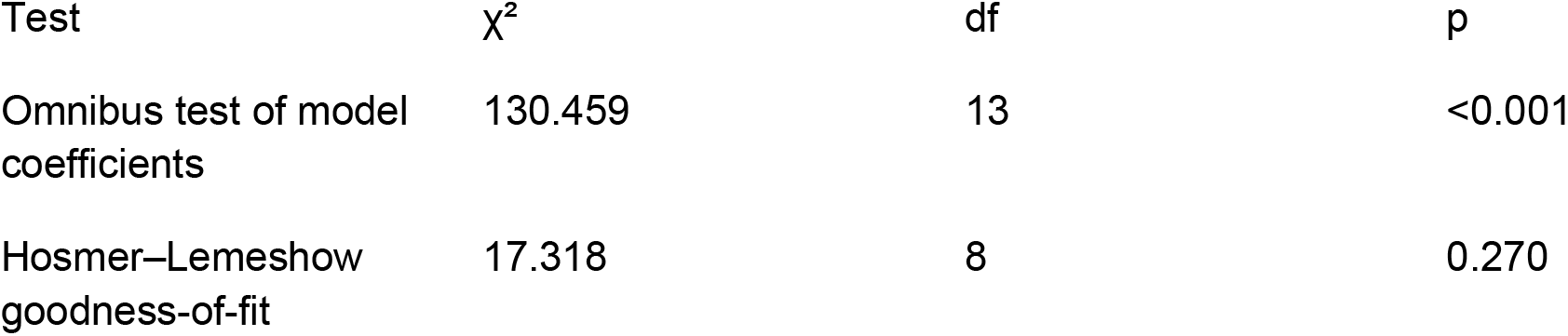

| Test | $\chi^2$ | df | p |
| --- | --- | --- | --- |
| Omnibus test of model coefficients | 130.459 | 13 | <0.001 |
| Hosmer–Lemeshow goodness-of-fit | 17.318 | 8 | 0.270 |

